# Contemporary Administrative Codes For Identifying Pulmonary Vein Isolation Procedures for Atrial Fibrillation

**DOI:** 10.1101/2024.02.12.24302143

**Authors:** Enrico G Ferro, Matthew R. Reynolds, Xu Jiaman, Yang Song, David J. Cohen, Rishi K Wadhera, Andre d’Avila, Peter J Zimetbaum, Robert W Yeh, Daniel B Kramer

**Author notes:** <u>Addresses for Correspondence</u>: Dr. Daniel B Kramer, TBD, 375 Longwood Ave, Boston, MA 02215. <u>Funding</u>: None. <u>Disclosures</u>: DJC reports institutional research grant support from Abbott, Boston Scientific, I-Rhythm, Edwards Lifescience, and Philips and consulting income from Abbott, Medtronic, Boston Scientific, and Edwards Lifesciences All other authors reported no relationships relevant to the contents of this paper to disclose.

## Abstract

**Introduction:** Use of pulmonary vein isolation (PVI) to treat atrial fibrillation continues to grow. Despite great interest in leveraging administrative data for real-world analyses, reliability of contemporary procedural codes for identifying PVI have not been carefully examined.

**Methods:** Inpatient PVIs were identified among US Medicare fee-for-service beneficiaries using Current Procedural Terminology (CPT) code 93656 in the Carrier Line Files. Each patient was cross-matched by procedure date with claims from Medicare Provider Analysis and Review Files (MedPAR) in order to compare CPT claims with International Classification of Diseases-10^th^ Revision Procedure Coding System (ICD-10-PCS) claims submitted by healthcare facilities to bill for the same procedure. We performed the reverse analysis for commonly matched ICD-10-PCS codes to identify their corresponding CPT-billed procedures. Lastly, we reviewed a random selection of 100 institutional cases for additional comparison of CPT and ICD-10-PCS assignation.

**Results:** We identified 25,617 inpatient PVIs from 1/2017 to 12/2021, of which 18,165 (71%) were linked to MedPAR by same-day procedure date. Of these, 16,672 (92%) were coded as ICD-10 02583ZZ “Destruction of Conduction Mechanism, Percutaneous Approach”, with lower use of other codes. The reverse process yielded heterogeneous results: among 75,003 procedures billed as ICD-10 02583ZZ, only 15,691 (21%) matched with CPT 93656 (PVI), as several other procedures were interchangeably billed under this same ICD-10 code. Institutional case review confirmed the greater specificity of CPT codes for identifying PVIs.

**Conclusions:** The ICD-10-PCS code most commonly associated with CPT-billed PVI procedures actually refers to ablation of the atrio-ventricular junction. Yet this same ICD-10-PCS code also matches with a wide range of other procedures distinct from PVI. We conclude that ICD-10-PCS codes alone are neither sensitive nor specific for identifying PVIs in claims. CPT codes should be used for health services research on this important procedure.

## Introduction

Atrial fibrillation (AF) poses a large and growing burden on public health.^1-4^ Treatment strategies for AF, including new technologies for catheter ablation, continue to evolve with a focus on early and sustained rhythm control to improve clinical outcomes and quality of life.^5-8^ Catheter ablation of AF focusing primarily on pulmonary vein isolation (PVI) is superior to anti-arrhythmic drugs for reducing the burden of AF, and is increasingly supported as first-line therapy in symptomatic patients.^5,9-11^ Rapid growth in the procedure has sparked interest in using administrative data to characterize real-world outcomes.^12,13^

Prior studies in this area have important limitations. Investigators leveraging Current Procedural Terminology (CPT) codes, which may capture both inpatient and outpatient cases, or ICD-10 Procedure Coding System (PCS) codes, which are only available for inpatient cases, may ascertain cases differently and thus draw different conclusions about procedural characteristics and outcomes^.13,14^ Inpatient versus outpatient assignation for PVI may be partially idiosyncratic based on institutional or patient insurance factors, imperfectly aligns with actual length of stay, and represents a limited and non-representative sample of all encounters.^12^ Thus, reliable and accurate characterization of PVIs in administrative data remains an important gap in health services research for this procedure.

This study therefore used comprehensive data from the Centers of Medicare and Medicaid Services to evaluate CPT and ICD-10-PCS claims codes for identifying PVIs. We paired this with institutional chart review to further explore the alignment of CPT and ICD-10-PCS codes for electrophysiology encounters.

## Methods

### Ethics Statement

This protocol was deemed exempt by the Institutional Review Board at Beth Israel Deaconess Medical Center. Due to the restrictions of the data use agreement with Medicare under which this analysis was performed, study data cannot be shared with outside investigators.

### Data Source

The Medicare Carrier Line Files, which contain records for 100% of fee-for-service (FFS) Medicare beneficiaries, were accessed through the Virtual Research Data Center and used to identify all procedural and outcome data from both inpatient and outpatient encounters. Inpatient and outpatient status were adjudicated using place of service codes 21 (inpatient hospital) and 22 (on-campus outpatient hospital) respectively, with outpatient status incorporating both observation stays and same-day discharge encounters. Patient encounters associated with place of service code 19 (off-campus-outpatient hospital) were excluded as it is unlikely that PVI procedures would be performed in these settings in contemporary clinical practice.

The databases were queried from January 1, 2017 to December 31, 2021. January 2017 was chosen as the start of the study period as this was the first full calendar year after the mandatory nationwide transition (on October 2015) from the International Classification of Diseases 9^th^ revision (ICD-9) to the ICD-10 Clinical Modification for diagnosis codes.

### Code Selection

All PVI procedures were first identified using the Category 1 Current Procedural Terminology (CPT) codes as the hypothesized gold standard, against which a list of ICD-10 Procedure Coding System (PCS) codes were compared. Both CPT and ICD-10 PCS codes were pre-specified based on literature review and clinical expertise, and are listed in **Table 1**.

**Table 1.** Claims Codes to Identify Electrophysiologic Procedures.

| CPT Code Number | CPT Code Description |
| --- | --- |
| 93650 | Intracardiac catheter ablation of atrioventricular node (AVJ) function |
| 93653 | Intracardiac catheter ablation of arrhythmogenic focus (supraventricular tachycardia [SVT], accessory atrioventricular connection, cavo-tricuspid isthmus [CTI] or single atrial focus) |
| 93655* | Intracardiac catheter ablation of discrete arrhythmia which is distinct from primary ablated mechanism |
| 93656 | Intracardiac catheter ablation of atrial fibrillation by pulmonary vein isolation (PVI) |
| 93657* | Additional catheter ablation for treatment of atrial fibrillation remaining after completion of pulmonary vein (PV) isolation |
| ICD Code Number | ICD-10 PCS Code Description |
| 02573ZZ | Destruction of Left Atrium (LA), Percutaneous Approach |
| 02583ZZ | Destruction of Conduction Mechanism, Percutaneous Approach |
| 025T3ZZ | Destruction of Right Pulmonary Vein (RPV), Percutaneous Approach |
| 025S3ZZ | Destruction of Left Pulmonary Vein (LPV), Percutaneous Approach |

### Study Population

All Medicare FFS beneficiaries aged 65 or older were eligible to enter the study cohort at the time of their index PVI, defined as the first recorded procedural claim, with a prior or concurrent diagnosis code for AF. We excluded patients who were enrolled in Medicare FFS for less than one year before index PVI, or those who had any prior AF-related catheter or surgical ablation codes (i.e. MAZE procedures) identified in the 3-year lookback period before their index PVI. Given the hypothesized high specificity of CPT codes in describing the PVI procedure and distinguishing it from other ablations for other supraventricular tachycardias, we did not exclude patients who carried concomitant diagnoses of other supraventricular arrythmias prior to or concurrent with their index PVI.

### Comparison of Claims Codes to Identify PVI Procedures

Inpatient PVI procedures were first identified using CPT code 93656 in the Carrier files as the gold standard – due to the fact that this CPT code describes the PVI procedure in more granular detail compared to various available ICD-10 PCS codes and has remained unchanged since 2012 (reducing the risk of heterogeneity in coding practices).

Each PVI identified on the Carrier files was then cross-matched by procedure date with claims from the Medicare Provider Analysis and Review (MedPAR) file. This is a separate database that contains information on inpatient hospitalizations (or in skilled nursing facilities), and can be used to identify inpatient procedures. Therefore, CPT claims submitted for PVI on the Carrier files were compared with ICD-10 PCS claims submitted by hospital sites on MedPAR to bill for each PVI. We then performed the reverse analysis for commonly matched ICD-10-PCS codes, to identify their corresponding CPT-billed procedures.

### Institutional Review

For additional assessment of our code selection and case ascertainment, we identified a random sample of 100 patients who underwent electrophysiology procedures at Beth Israel Deaconess Medical Center (Boston, MA) coded as either “inpatient” or “outpatient” between 06/01/2016 and 06/01/2022. Patient charts were identified by querying the electronic medical record for the CPT and ICD-10 PCS procedural codes of interest. Manual chart review of the electrophysiology procedure report was performed by E.G.F and D.B.K. to adjudicate which procedures were performed in each case and confirm whether the billing codes ultimately matched the performed procedure, based on clinical expertise.

All analyses were conducted in SAS version 9.4 (SAS Institute, Cary, North Carolina), with a p-value less than 0.05 considered to represent statistical significance.

## Results

### Identification and Code Comparison of PVI Procedures

Between January 2017 and December 2021, a total of 205,887 Medicare beneficiaries with AF underwent first-time PVI. From this initial sample, we identified a total of 25,617 inpatient PVIs (based on CPT code 93656) in the Carrier files (**Figure 1**). Among those, 18,165 (70.9%) were successfully linked to MedPAR by same-day procedure date. Of these linked procedures, 16,672 (91.7%) were billed as ICD-10 PCS 02583ZZ “*Destruction of Conduction Mechanism, Percutaneous Approach*”, with less frequent and fragmented use of other codes in the remaining 8.3% of cases (**Figure 1**).

**Figure 1.**
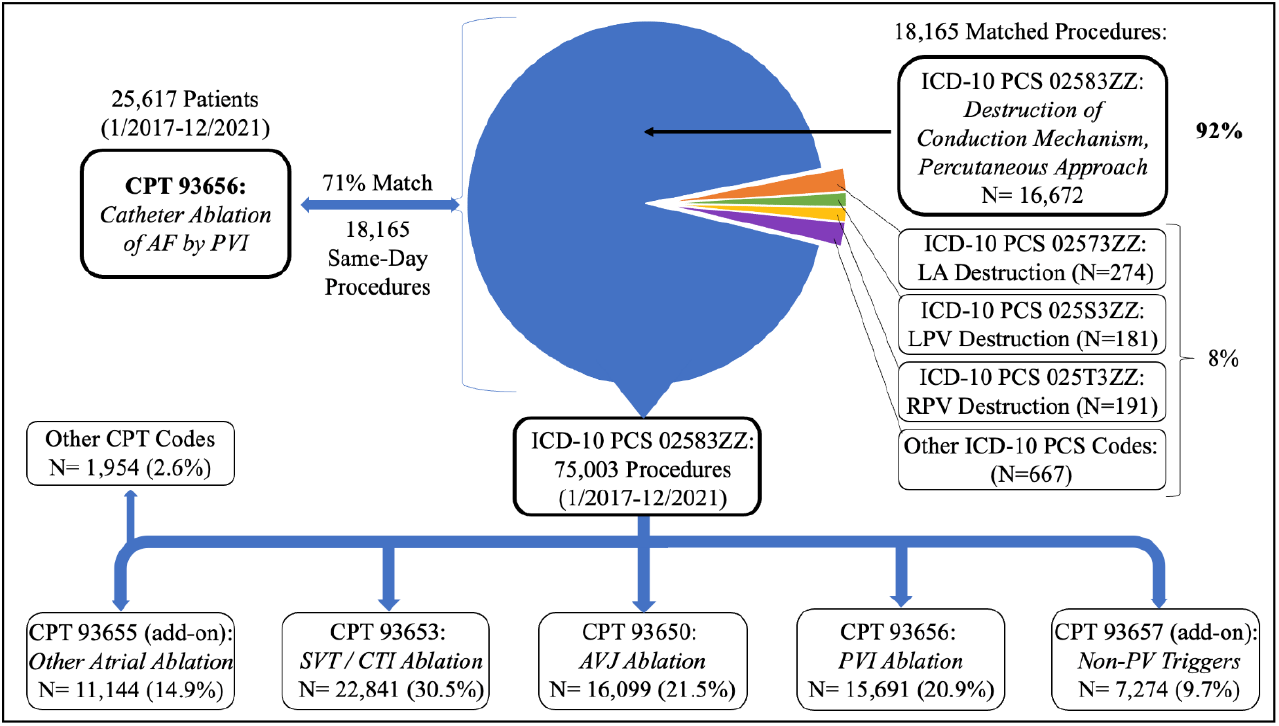
Cross-Validation of ICD-10-PCS and CPT Codes to Identify PVI Procedures. Top: Mapping of CPT-adjudicated PVI (ground truth) to ICD-10 PCS claims with same procedure date. Bottom: Mapping of most commonly used ICD-10-PCS code (02583ZZ) to corresponding procedures with same date by CPT claims. AF, atrial fibrillation; AVJ, atrioventricular junction; CPT, current procedural terminology code; CTI, cavotricuspid isthmus ablation; ICD-10 PCS, International Classification of Diseases 10th Revision, Procedure Coding System; LA, left atrium; LPV, left pulmonary vein; RPV, right pulmonary vein; PVI, pulmonary vein isolation; SVT, supraventricular tachycardia.

The reverse process yielded heterogeneous results. In this case, we started the claim validation process from ICD-10 PCS 02583ZZ, since this was found to be the most commonly used ICD-10 code that matched with the CPT code 93656 in the Carrier files. Among 75,003 procedures billed as ICD-10 PCS 02583ZZ in the MedPAR files, only 15,691 (20.9%) matched with CPT 93656 in the Carrier files by same-day procedure date. Several other procedures were interchangeably billed with a heterogeneous list of CPT codes, all of which cross-matched to the same ICD-10 PCS 02583ZZ code (**Figure 1**).

### Institutional Review

A similar pattern was identified through manual chart review of electrophysiology reports from a random sample of 100 patients who underwent a variety of procedures at Beth Israel Deaconess Medical Center in both inpatient and outpatient settings between 06/01/2016 and 06/01/2022 **(Table 2)**. Specifically, ICD-10 PCS 02583ZZ was used in virtually all inpatient procedures, even though none of these procedures involved ablation of the conduction mechanism (i.e. the atrioventricular junction). Other ICD-10 PCS codes were used in very few cases, such as 02573ZZ “*Destruction of Left Atrium, Percutaneous Approach*”. In these cases, the alternative ICD-10 PCS codes matched with the clinical description of the procedure provided in the electrophysiology report – such as ablation of a left-sided atrial flutter.

**Table 2.** Validation of Claims Codes with Retrospective Chart Review.

| **Fiscal Year** | **Patient Status** | **ICD-10-PCS Code** | **CPT Code** | **Procedure Performed \*** | **Clinical Agreement with ICD-10 PCS** | **Clinical Agreement with CPT** |
| --- | --- | --- | --- | --- | --- | --- |
| 2016 | Inpatient | 02583ZZ | 93656 | PVI | No | Yes |
| 2016 | Inpatient | 02583ZZ | 93563 | CTI | No | Yes |
| 2016 | Inpatient | 02583ZZ | 93653 | CTI | No | Yes |
| 2016 | Inpatient | 02573ZZ | 93653 | SVT | Yes | Yes |
| 2016 | Inpatient | 02583ZZ | 93653 | CTI | No | Yes |
| 2016 | Inpatient | 02583ZZ | 93655, 93656 | PVI, CTI | No | Yes |
| 2016 | Inpatient | 02583ZZ | 93653 | SVT | No | Yes |
| 2016 | Inpatient | 02583ZZ | 93653, 93656 | PVI, CTI | No | Yes |
| 2016 | Inpatient | 02583ZZ | 93655 | SVT | No | Yes |
| 2016 | Inpatient | 025S3ZZ | 93656 | SVT | No | No |
| 2016 | Inpatient | 02583ZZ | 93653 | SVT | No | Yes |
| 2016 | Inpatient | 02583ZZ | 93653 | CTI | No | Yes |
| 2016 | Inpatient | 02583ZZ | 93653 | CTI | No | Yes |
| 2016 | Inpatient | 02583ZZ | 93656 | PVI | No | Yes |
| 2016 | Inpatient | 02583ZZ | 93653 | SVT | No | Yes |
| 2016 | Inpatient | 02583ZZ | 93656 | PVI | No | Yes |
| 2016 | Inpatient | 02583ZZ | 93653 | CTI | No | Yes |
| 2016 | Inpatient | 02583ZZ | 93655, 93656 | PVI, PWI | No | Yes |
| 2016 | Inpatient | 02583ZZ | 93655, 93656 | PVI, CTI | No | Yes |
| 2017 | Inpatient | 02583ZZ | 93656 | PVI | No | Yes |
| 2017 | Inpatient | 02583ZZ | 93653, 93655 | SVT, CTI | No | Yes |
| 2017 | Inpatient | 02583ZZ | 93656 | PVI | No | Yes |
| 2017 | Inpatient | 02573ZZ | 93655, 93656 | PVI, PWI | Yes | Yes |
| 2017 | Inpatient | 02583ZZ | 93656 | PVI | No | Yes |
| 2017 | Inpatient | 02583ZZ | 93656 | PVI | No | Yes |
| 2017 | Inpatient | 02583ZZ | 93656 | PVI | No | Yes |
| 2018 | Inpatient | 02583ZZ | 93656 | PVI | No | Yes |
| 2018 | Inpatient | 02583ZZ, 025T3ZZ | 93656 | PVI | Yes | Yes |
| 2018 | Inpatient | 02583ZZ | 93655, 93656 | PVI, CTI | No | Yes |
| 2018 | Inpatient | 02583ZZ | 93656 | PVI | No | Yes |
| 2018 | Inpatient | 02583ZZ | 93655, 93656 | PVI, SVT | No | Yes |
| 2018 | Inpatient | 02583ZZ | 93655, 93656 | PVI, CTI | No | Yes |
| 2019 | Inpatient | 02583ZZ | 93653 | CTI | No | Yes |
| 2019 | Inpatient | 02583ZZ | 93653, 93655 | SVT | No | Yes |
| 2019 | Inpatient | 02583ZZ | 93655, 93656 | PVI, CTI | No | Yes |
| 2019 | Inpatient | 02583ZZ | 93655, 93656 | PVI, CTI | No | Yes |
| 2019 | Inpatient | 02583ZZ | 93655, 93656 | PVI, CTI | No | Yes |
| 2019 | Inpatient | 02573ZZ | 93653 | SVT | No | Yes |
| 2020 | Inpatient | 02583ZZ | 93653, 93655 | SVT | No | Yes |
| 2020 | Inpatient | 02583ZZ | 93653, 93655 | SVT, CTI | No | Yes |
| 2020 | Inpatient | 025S3ZZ, 025T3ZZ | 93655, 93656 | PVI, CTI | Yes | Yes |
| 2020 | Inpatient | 02583ZZ | 93655, 93656 | PVI, CTI | No | Yes |
| 2020 | Inpatient | 02583ZZ | 93655, 93656 | PVI, PWI | No | Yes |
| 2021 | Inpatient | 02573ZZ, 025S3ZZ<br>025T3ZZ | 93656 | PVI | Yes | Yes |
| 2021 | Inpatient | 02583ZZ | 93655, 93656 | PVI, CTI | No | Yes |
| 2021 | Inpatient | 02583ZZ | 93655, 93656 | PVI, PWI | No | Yes |
| 2021 | Inpatient | 02583ZZ | 93655, 93656 | PVI, CTI | No | Yes |
| 2021 | Inpatient | 02583ZZ | 93655, 93656 | PVI, PWI | No | Yes |
| 2021 | Inpatient | 02583ZZ | 93655, 93656 | PVI, CTI | No | Yes |
| 2021 | Inpatient | 02583ZZ | 93655, 93656 | PVI, CTI | No | Yes |
| 2021 | Inpatient | 02583ZZ | 93655, 93656 | PVI, PWI | No | Yes |
| 2021 | Inpatient | 02583ZZ | 93655, 93656 | PVI, PWI | No | Yes |
| 2021 | Inpatient | 02583ZZ | 93655, 93656 | PVI, SVT | No | Yes |
| 2021 | Inpatient | 02583ZZ | 93655, 93656 | PVI, CTI | No | Yes |
| 2022 | Inpatient | 02583ZZ | 93653, 93655 | CTI | No | Yes |
| 2022 | Inpatient | 02583ZZ | 93655, 93656 | PVI, CTI | No | Yes |
| 2022 | Inpatient | 02583ZZ | 93653 | CTI | No | Yes |
| 2022 | Inpatient | 02583ZZ | 93655, 93656 | PVI, CTI | No | Yes |
| 2016 | Outpatient | N/A | 93655, 93656 | PVI, SVT | N/A | Yes |
| 2016 | Outpatient | N/A | 93656 | PVI | N/A | Yes |
| 2016 | Outpatient | N/A | 93653 | SVT | N/A | Yes |
| 2016 | Outpatient | N/A | 93653 | CTI | N/A | Yes |
| 2016 | Outpatient | N/A | 93655, 93656 | PVI, PWI | N/A | Yes |
| 2016 | Outpatient | N/A | 93656 | PVI | N/A | Yes |
| 2016 | Outpatient | N/A | 93656 | PVI | N/A | Yes |
| 2016 | Outpatient | N/A | 93653 | CTI | N/A | Yes |
| 2016 | Outpatient | N/A | 93653 | SVT | N/A | Yes |
| 2017 | Outpatient | N/A | 93656 | PVI | N/A | Yes |
| 2017 | Outpatient | N/A | 93655, 93656 | PVI, PWI | N/A | Yes |
| 2017 | Outpatient | N/A | 93653, 93655 | SVT | N/A | Yes |
| 2017 | Outpatient | N/A | 93655, 93656, 93657 | PVI, PWI, ML | N/A | Yes |
| 2018 | Outpatient | N/A | 93655, 93656 | PVI, CTI | N/A | Yes |
| 2018 | Outpatient | N/A | 93653, 93655 | SVT, CTI | N/A | Yes |
| 2018 | Outpatient | N/A | 93653 | SVT | N/A | Yes |
| 2018 | Outpatient | N/A | 93655, 93656 | PVI, SVT | N/A | Yes |
| 2018 | Outpatient | N/A | 93653 | SVT | N/A | Yes |
| 2018 | Outpatient | N/A | 93656 | SVT | N/A | Yes |
| 2018 | Outpatient | N/A | 93655, 93656 | PVI, CTI | N/A | Yes |
| 2019 | Outpatient | N/A | 93656 | SVT | N/A | No |
| 2019 | Outpatient | N/A | 93655, 93656 | PVI, CTI | N/A | Yes |
| 2019 | Outpatient | N/A | 93655, 93656 | PVI, CTI | N/A | Yes |
| 2019 | Outpatient | N/A | 93653 | SVT | N/A | Yes |
| 2019 | Outpatient | N/A | 93653 | SVT | N/A | Yes |
| 2019 | Outpatient | N/A | 93655, 93656 | PVI, SVT | N/A | Yes |
| 2019 | Outpatient | N/A | 93655, 93656 | PVI, CTI | N/A | Yes |
| 2020 | Outpatient | N/A | 93653, 93655 | SVT | N/A | Yes |
| 2020 | Outpatient | N/A | 93655, 93656 | PVI, CTI | N/A | Yes |
| 2020 | Outpatient | N/A | 93653, 93655 | SVT | N/A | Yes |
| 2021 | Outpatient | N/A | 93655, 93656 | PVI, CTI | N/A | Yes |
| 2021 | Outpatient | N/A | 93653 | PVI | N/A | No |
| 2021 | Outpatient | N/A | 93653 | SVT | N/A | Yes |
| 2021 | Outpatient | N/A | 93653, 93655 | SVT | N/A | Yes |
| 2021 | Outpatient | N/A | 93655, 93656 | PVI, SVT | N/A | Yes |
| 2021 | Outpatient | N/A | 93655, 93656 | PVI, CTI | N/A | Yes |
| 2021 | Outpatient | N/A | 93655, 93656 | PVI, CTI | N/A | Yes |
| 2021 | Outpatient | N/A | 93655, 93656 | PVI, CTI | N/A | Yes |
| 2021 | Outpatient | N/A | 93655, 93656, 93657 | PVI, PWI, ML | N/A | Yes |
| 2021 | Outpatient | N/A | 93655, 93656,<br>93657 | PVI, PWI,<br>ML | N/A | Yes |
| 2021 | Outpatient | N/A | 93655, 93656 | PVI, CTI | N/A | Yes |
| 2022 | Outpatient | N/A | 93653,<br>93655 | SVT, CTI | N/A | Yes |
| 2022 | Outpatient | N/A | 93653 | SVT | N/A | Yes |

Among outpatient procedures, there was nearly universal match between the CPT code used for billing purposes and the clinical description of the procedure provided in the electrophysiology report, whether PVI or catheter ablation of other supraventricular tachycardias distinct from AF.

## Discussion

This study clarifies the importance of using comprehensive codes to accurately ascertain contemporary PVI volumes and outcomes using claims-based data. When cross-matching ICD-10-PCS and CPT codes, we found that the ICD-10-PCS code most commonly billed for PVI procedures actually refers to ablation of the atrioventricular junction (AVJ) – which should be excluded when designing analyses to study AF ablation, based on the clinical difference between the two procedures and their target populations. AVJ ablation is typically reserved for older, higher-risk patients as a last resort to manage permanent AF. Furthermore, the ICD-10-PCS code for AVJ ablation was also found to cross-match with CPT codes for a wide range of ablations that are unrelated to PVI. Notably, we confirmed these findings from the Medicare database in the internal medical record of our institution - where we could perform a more meticulous clinical review of each procedure report. Our findings thus show that ICD-10-PCS codes alone are neither sensitive nor specific for identifying PVI in claims, and cannot reliably be used for clinical outcomes research related to this procedure.^15,16^

The claims comparison exercise performed in our study suggests that CPT codes should be systematically prioritized and adopted to reliably identify PVI, as already demonstrated for other cardiovascular procedures.^15,17^,18 The accuracy of this approach most likely results from the fact that CPT codes describe the procedure of interest in more detail, while ICD-10-PCS codes offer shorter and vaguer descriptions that leave room for interpretation and resulting heterogeneity in coding practices. Moreover, in AF ablation as in other procedures, CPT codes tend to remain unchanged across several years, while ICD codes go through multiple iterations that limit their stability over time. Lastly, CPT codes are more often selected by the operators who perform the procedure, and less often by hospital billing departments, who may select ICD-10-PCS codes based on limited understanding and interpretation of procedural notes.

Our analysis has several limitations. This Medicare-based analysis is limited to patients older than 65 years and does not capture information on ablations performed on younger patients or those with other insurance. Claims-based analyses cannot provide granular information on valuable technical variables such as thermal sources (i.e. radiofrequency vs cryoballoon), ablation catheters or peri-procedural imaging. Our current analysis is limited to validation of the PVI administrative codes, yet other lesion sets may be performed during an AF procedure (e.g., posterior wall isolation, mitral lines, etc), which may influence procedural outcomes. However, as our study was limited to first-time AF ablations, we suspect that the incidence of non-PVI lesion sets is small.

## Conclusions

We found that CPT codes should be systematically adopted to reliably identify PVI in administrative claims data, since ICD-10-PCS codes alone are neither sensitive nor specific. As claims-based research increases in popularity, this must be matched by an equally rigorous understanding of its methodological nuances.^19^

## Data Availability

Due to the restrictions of the data use agreement with Medicare under which this analysis was performed, study data cannot be shared with outside investigators.

## Acknowledgements

None

## References

1. Hindricks G, Potpara T, Dagres N, et al. 2020 ESC Guidelines for the diagnosis and management of atrial fibrillation developed in collaboration with the European Association for Cardio-Thoracic Surgery (EACTS): The Task Force for the diagnosis and management of atrial fibrillation of the European Society of Cardiology (ESC) Developed with the special contribution of the European Heart Rhythm Association (EHRA) of the ESC. Eur Heart J. 2021;42(5):373–498.

2. January CT, Wann LS, Calkins H, et al. 2019 AHA/ACC/HRS Focused Update of the 2014 AHA/ACC/HRS Guideline for the Management of Patients With Atrial Fibrillation: A Report of the American College of Cardiology/American Heart Association Task Force on Clinical Practice Guidelines and the Heart Rhythm Society in Collaboration With the Society of Thoracic Surgeons. Circulation. 2019;140(2):e125–e151.

3. Chugh SS, Havmoeller R, Narayanan K, et al. Worldwide epidemiology of atrial fibrillation: a Global Burden of Disease 2010 Study. Circulation. 2014;129(8):837–847.

4. Colilla S, Crow A, Petkun W, Singer DE, Simon T, Liu X. Estimates of current and future incidence and prevalence of atrial fibrillation in the U.S. adult population. The American journal of cardiology. 2013;112(8):1142–1147.

5. Camm AJ, Naccarelli GV, Mittal S, et al. The Increasing Role of Rhythm Control in Patients With Atrial Fibrillation: JACC State-of-the-Art Review. J Am Coll Cardiol. 2022;79(19):1932–1948.

6. Kirchhof P, Camm AJ, Goette A, et al. Early Rhythm-Control Therapy in Patients with Atrial Fibrillation. N Engl J Med. 2020;383(14):1305–1316.

7. Marrouche NF, Brachmann J, Andresen D, et al. Catheter Ablation for Atrial Fibrillation with Heart Failure. N Engl J Med. 2018;378(5):417–427.

8. Mark DB, Anstrom KJ, Sheng S, et al. Effect of Catheter Ablation vs Medical Therapy on Quality of Life Among Patients With Atrial Fibrillation: The CABANA Randomized Clinical Trial. JAMA. 2019;321(13):1275–1285.

9. Wazni OM, Dandamudi G, Sood N, et al. Cryoballoon Ablation as Initial Therapy for Atrial Fibrillation. N Engl J Med. 2021;384(4):316–324.

10. Kuck KH, Lebedev DS, Mikhaylov EN, et al. Catheter ablation or medical therapy to delay progression of atrial fibrillation: the randomized controlled atrial fibrillation progression trial (ATTEST). Europace. 2021;23(3):362–369.

11. Writing Committee M, Joglar JA, Chung MK, et al. 2023 ACC/AHA/ACCP/HRS Guideline for the Diagnosis and Management of Atrial Fibrillation: A Report of the American College of Cardiology/American Heart Association Joint Committee on Clinical Practice Guidelines. J Am Coll Cardiol. 2023.

12. Reynolds MR, Kramer DB, Yeh RW, Cohen DJ. AF Ablation Outcomes: Real World or Fun House Mirror? J Am Coll Cardiol. 2020;75(10):1243.

13. Cheng EP, Yeo I, Kim LK, Lerman BB, Cheung JW. Reply: Early Mortality Following Catheter Ablation of Atrial Fibrillation. J Am Coll Cardiol. 2020;75(10):1245–1247.

14. Loring Z, Holmes DN, Matsouaka RA, et al. Procedural Patterns and Safety of Atrial Fibrillation Ablation: Findings From Get With The Guidelines-Atrial Fibrillation. Circ Arrhythm Electrophysiol. 2020;13(9):e007944.

15. Ferro EG, Kramer DB, Li S, et al. Incidence, Treatment, and Outcomes of Symptomatic Device Lead-Related Venous Obstruction. J Am Coll Cardiol. 2023;81(24):2328–2340.

16. Raja A, Dicks A, Sardar P, Schermerhorn ML, Yeh RW, Secemsky EA. Accuracy of Administrative Claims Codes for Identifying Devices Used in Endovascular Femoropopliteal Artery Revascularisation: A Retrospective Observational Study at Two Tertiary Centres in the United States. Eur J Vasc Endovasc Surg. 2022;63(5):769–770.

17. Secemsky EA, Raja A, Shen C, Valsdottir LR, Schermerhorn M, Yeh RW. Rationale and Design of the SAFE-PAD Study. Circulation: Cardiovascular Quality and Outcomes. 2021;14(1):e007040.

18. Secemsky EA, Shen C, Schermerhorn M, Yeh RW. Longitudinal Assessment of Safety of Femoropopliteal Endovascular Treatment With Paclitaxel-Coated Devices Among Medicare Beneficiaries: The SAFE-PAD Study. JAMA Internal Medicine. 2021;181(8):1071–1080.

19. Yeh RW. Bringing the Credibility Revolution to Observational Research in Cardiology. Circulation. 2023;148(6):455–458.

